# Frequent lncRNA-derived fusions in pediatric neuroblastoma identified by *LncFusion*: potential biomarker and therapeutic implications

**DOI:** 10.1101/2025.01.16.25320696

**Authors:** Zixiu Li, Joae Wu, Peng Zhou, Chan Zhou

## Abstract

Despite growing interest in the onco-fusion proteins and long noncoding RNAs (lncRNAs) in cancers, lncRNA-derived fusion transcripts in pediatric cancers remain understudied. To address this gap, we first developed *LncFusion*, a novel computational pipeline that systematically detects lncRNA-derived fusion transcripts from RNA-seq data. Leveraging our previously published *Flnc* (for comprehensive lncRNA identification) as a foundation and applying *LncFusion*, we identified over 900 high-confidence lncRNA-derived fusions (lnc-fusions) in pediatric neuroblastoma by analyzing the transcriptomics datasets from three major pediatric cancer cohorts—TARGET, Gabriella Miller Kids First, and St. Jude Cloud. The number of lnc-fusions exceeds the number of mRNA-derived fusions (mRNA-fusions) in neuroblastoma. Whole genome sequencing analyses revealed that approximately 40% of these lnc-fusions result from chromosomal rearrangements, while over 60% may arise from aberrant splicing events. Among these high-confidence lnc-fusions, 61 are enriched in pediatric neuroblastoma compared to healthy controls; and 20s exhibit subtype-specific expressions in pediatric neuroblastoma patients, which would be categorized into three groups: MYCN-amplified patients, c-MYC-highly expressed patients and the remaining (MYCN-unamplified and c-MYC not high expression). Subtype-specific enrichment of certain lnc-fusions, particularly in MYCN-amplified and c-MYC-high subgroups, underscores distinct oncogenic roles. Further functional studies implicated lnc-fusions in key pathways related to neuron development, translation, and energy metabolism, suggesting potential contributions to neuroblastoma pathogenesis. Additionally, We found several novel fusions might serve as potential diagnostic or prognostic biomarkers in neuroblastoma. A few candidates correlate with either favorable histology and lower-risk patient subsets, or poorer survival outcomes, indicating strong prognostic biomarker potential. Experiments in cell line further confirmed the presence of a few key lnc-fusions discovered from patient samples. Our findings provide the first comprehensive insight into lncRNA-derived fusions in pediatric neuroblastoma, providing the promise of lnc-fusions as novel biomarkers and therapeutic targets. The *LncFusion* tool developed can also be applied to explore lnc-fusions in other pediatric and adult cancers.

## INTRODUCTION

Pediatric cancer remains a significant global health concern and is the leading cause of disease-related death among children^1,2^. Unlike many adult cancers characterized by a high genetic mutation rate, pediatric cancers often harbor fewer genetic mutations^3–5^. Instead, fusion transcripts and their protein products are common in pediatric cancer compared to adult cancers^3–5^. Oncogenic fusions are increasingly recognized as key drivers of pediatric tumorigenesis^6^ and attractive targets for biomarker and therapy development^6,7^. Several fusion transcripts and their protein products have been exploited clinically in pediatric cancer. For example, the EWS-FLI1 and PAX3-FOX01 fusions are used to assist diagnosis and guide treatment in pediatric sarcomas children ^6,8,9^. Despite these notable examples, many fusion events in childhood cancers remain unexplored, particularly those involving long noncoding RNAs (lncRNAs).

Long noncoding RNAs (lncRNAs) are long linear transcripts that generally do not encode proteins, typically ranging from 1,000 to 10,000 bp with a minimum length of 200 bp. But they play crucial roles in gene regulation, modulate cellular processes and disease progression^10–19^. lncRNAs outnumber protein-coding message RNAs (mRNAs) and often exhibit tissue- and disease-specific expression^20^. While thousands of lncRNA-derived fusion transcripts (lnc-fusions) have been identified in adult tumors^21^, their existence and significance in pediatric cancers remain largely unknown. Moreover, many pediatric-specific lncRNAs are unannotated, making it difficult for existing fusion-detection pipelines—which rely heavily on known gene annotations—to systematically capture these novel fusion transcripts.

To address these gaps, we developed *LncFusion*, a computational pipeline that systematically detects lncRNA-derived fusion transcripts (including annotated and unannotated lncRNAs) from large-scale RNA-sequencing (RNA-seq) data. *LncFusion* leverages our *de novo* lncRNA identification method, *Flnc*^22^, and rigorous filtering to discover high-confidence fusion events that may otherwise escape detection by conventional tools designed primarily for mRNA-derived fusions (mRNA-fusions).

In this study, we applied *LncFusion* to pediatric neuroblastoma, which is the most common malignant (cancerous) extracranial solid tumor of childhood, arising from tissues that form the sympathetic nervous system^23,24^. Neuroblastoma exhibits wide clinical heterogeneity, ranging from spontaneously regressing tumors to aggressive, treatment-resistant disease^25^. While prior investigations have identified mRNA-derived fusions in neuroblastoma^26–28^, systematic exploration of lnc-fusions in this cancer type is still lacking.

Here, by analyzing multiple neuroblastoma cohorts from major pediatric consortia (e.g., TARGET, Gabriella Miller Kids First, and St. Jude Cloud), we discovered over 900 high-confidence lnc-fusions. Many lnc-fusions frequently occur in pediatric neuroblastoma patients and are over-expressed in tumors compared to healthy controls. Some lnc-fusions exhibit subtype specificity (e.g., *MYCN*-amplified vs. non-amplified tumors) and potential roles in oncogenic pathways. We also demonstrated that a subset of these lnc-fusions correlates with clinical outcomes, suggesting their value as diagnostic or prognostic biomarkers. Additionally, we explored whether the identified lnc-fusions arise via chromosomal rearrangements or aberrant splicing^29,30^, providing deeper insight into their genetic underpinnings.

Overall, this work highlights a previously under-appreciated class of fusion transcripts in pediatric neuroblastoma and underscores the importance of integrating lncRNA-derived fusions through our *LncFusion* pipeline into pediatric oncology research. By uncovering lnc-fusions that may serve as potential biomarkers or therapeutic targets, our findings open new avenues for precision medicine approaches in childhood cancers.

## MATERIALS AND METHODS

### Patient cohorts and datasets collection

RNA-seq datasets of neuroblastoma (NB) patients were collected from the TARGET project (dbGaP acc: phs000218), the Gabriella Miller Kids First Pediatric Research Program (dbGaP acc: phs001228], and the St. Jude Cloud pediatric cancer omics data sharing ecosystem^31^. Specifically, 162 polyA-selected RNA-seq samples from TARGET, 182 Ribominus RNA-seq and 229 WGS samples from Kids First, and 171 Ribominus RNA-seq samples from St. Jude Cloud were obtained. Additionally, 258 polyA-selected RNA-seq datasets from normal adrenal gland tissues in the GTEx database^32^ and 39 neuroblastoma cell line RNA-seq datasets^33^ were included.

### Identification of lncRNAs

We used polyA-selected RNA-seq data (162 TARGET neuroblastoma samples + 258 GTEx normal adrenal gland samples) to identify novel and annotated lncRNAs with *Flnc*^22^.

### Fusion detection pipeline (*LncFusion*)

We developed *LncFusion* to identify lncRNA-involved fusions (lnc-fusions) and mRNA– mRNA fusions. A comprehensive index of lncRNA and mRNA was built by merging LncBook [citation], GENCODE (v42)^34^, and novel lncRNAs identified by Flnc. Three fusion callers— STAR-Fusion^35^, Arriba^36^, and STAR-SEQR^35^—were used. Putative fusions were retained if detected by at least two tools or met an FFPM ≥ 0.1 for STAR-Fusion-only calls. Mitochondrial, immunoglobulin, or highly duplicated genes were removed^37^. Fusions were classified as lnc-fusions (lncRNA–lncRNA or lncRNA–mRNA) or mRNA-fusions (mRNA–mRNA). *Please refer to the Supplemental Materials and Methods for the detailed description*.

### Visualization of Supporting Reads

We employed FusionInspector^38^ to visualize read alignments anchoring each breakpoint. This tool reconstructs candidate fusion transcripts, showing split/spanning read evidence to confirm or refute each fusion event.

### Whole-Genome Sequencing (WGS) date analysis

Breakpoints were validated in matched WGS data from 182 patients of Kids First cohort by counting spanning reads (MAPQ ≥ 20) and split reads (MAPQ ≥ 20). Spanning reads are paired-end reads in which one mate aligns to one gene, while the other mate aligns to a different gene. Split reads (also referred to as “chimeric reads”) more precisely localize fusion junctions by spanning both sides of the breakpoint within a single read. One read segment aligns to a gene, while the remaining segment aligns to a different gene. A fusion was supported if at least three reads, including spanning and split reads anchored the rearrangement, and each breakpoint was within 100 kb of the corresponding fusion partner^37^.

### Neuroblastoma subtype classification

We used MYCN and c-MYC expression, corrected for batch effects by ComBat^39^ after quantile normalization by limma package^40^, for KNN clustering (K=3). Subtypes were defined as (1) MYCN-amplified, (2) c-MYC-High and MYCN-unamplified, and (3) c-MYC-Low and MYCN-unamplified^41^.

### Differential expression analysis

Differentially expressed fusions were identified using a one-sided Wilcoxon rank-sum test^42^ with BH correction and fold change threshold. Comparisons were made to normal adrenal tissues (GTEx) and among neuroblasoma subtypes. Fusions with adjusted p-value < 0.05 and ≥2-fold change were considered significantly upregulated in neuroblastoma.

### Survival analysis

Overall survival data from TARGET and Kids First cohorts were analyzed using Kaplan– Meier curves and log-rank tests. P-values were derived from a chi-square distribution.

### Clinical associations

Associations between fusion status and clinical parameters (age, gender, tumor location, and disease severity) were tested using the Wilcoxon rank-sum test or Fisher’s exact test, as appropriate.

### Co-expression network and GO enrichment analysis

We constructed co-expression networks (Spearman ≥ 0.4; p-value<1e-20) for fusion genes (FFPM) and protein-coding genes (FPKM) across 773 samples using the MCL-edge “mcxarray” program^43^ and visualized sub-networks in Cytoscape^44^. GO enrichment analysis was performed using the DAVID Functional Annotation platform (http://david.abcc.ncifcrf.gov)^45,46^ with the background set of protein-coding genes (FPKM>1 in ≥1 sample).

### Cell culture and RT-PCR experiments

LA-N-5 and SH-SY5Y cells were maintained in RPMI1640 medium supplemented with 10% FBS. Total RNA was extracted using Trizol, and fusion transcripts were amplified by PCR (HotStarTaq Master Mix) using primers that span the putative breakpoint. PCR products were electrophoresed on a 2% agarose gel. Primers targeting the head and tail genes of each candidate fusion are shown as follows:

XLOC_043415-URI1-for: 5’-CCTGCGTTCAGGTGATTCT-3’

XLOC_043415-URI1-rev: 5’-CTGCTTTGCTGAGCACTTTG-3’

## RESULTS

### Computational workflow of LncFusion

To systematically identify lncRNA-derived fusion transcripts (lnc-fusions) (Fig. 1A) in pediatric cancers, we developed the *LncFusion* computational pipeline. The workflow integrates annotations from novel lncRNAs identified by *Flnc*^22^, annotated lncRNAs from the LncBook database, and coding genes from the GENCODE database to build a comprehensive library of reference annotations (Fig. 1B). This step ensures that both annotated and unannotated lncRNAs expressed in the tissue of interest are captured. Breakpoint detection was carried out using three complementary tools— STAR-Fusion^35^, Arriba^36^, and STAR-SEQR^35^—selected for their speed and accuracy in cancer transcriptome analysis^47^ (Fig. 1B). A consensus approach was used to retain only fusions detected by at least two tools or those with an expression threshold above FFPM>0.1. Additional filtering steps excluded fusions involving mitochondrial, immunoglobulin, duplicated, or paralogous genes, as these are often false positives. This rigorous workflow results in high-confidence fusion events, including both lnc-fusions and mRNA-derived fusions (mRNA-fusions). The integration of these stringent criteria ensures robustness and reliability in fusion detection, addressing gaps in conventional pipelines that often overlook lnc-fusions. Please refer to the Materials and Methods for details about the *LncFusion*.

**Figure 1.**
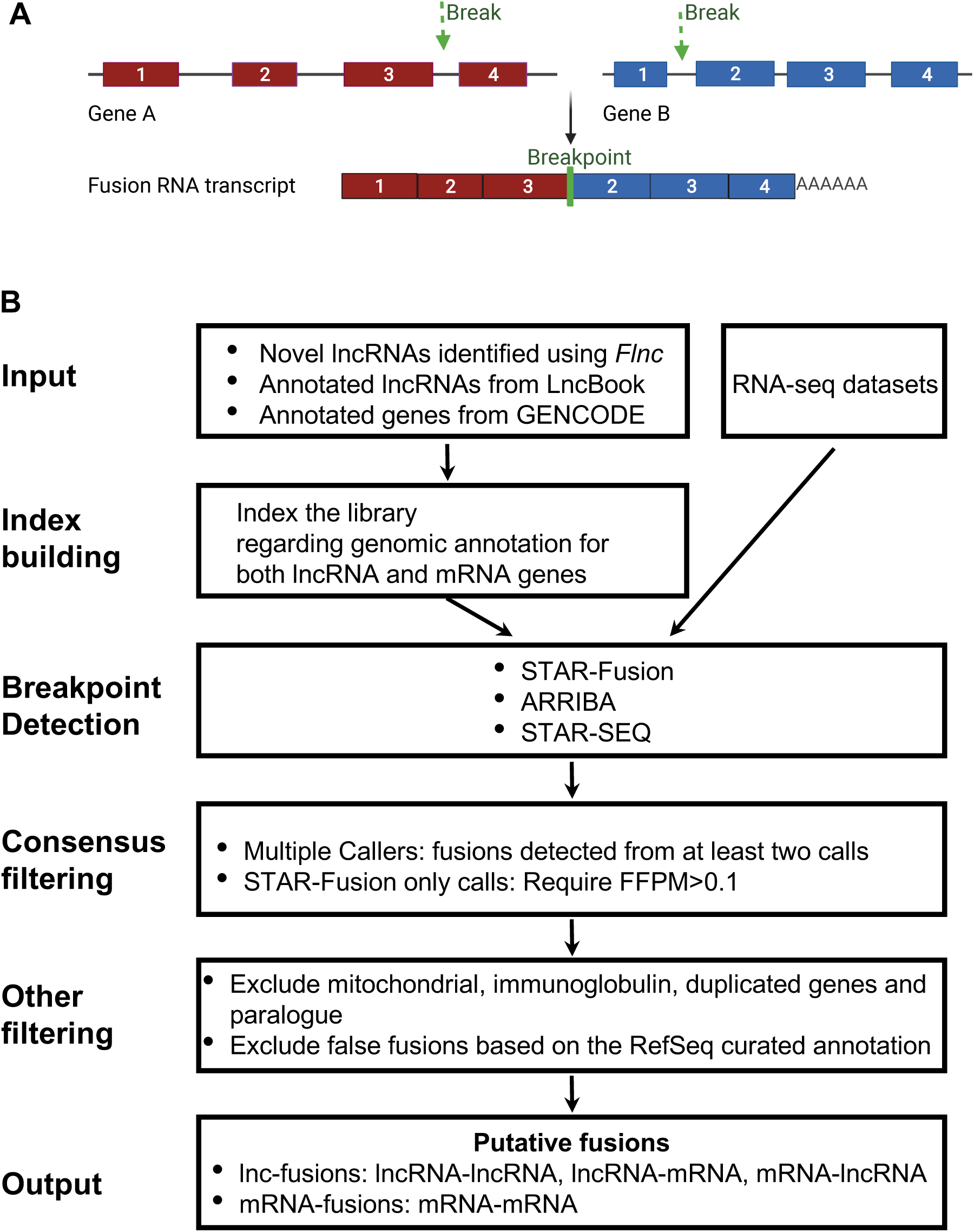
Computational workflow of *LncFusion*. (A) Schematic representation of a fusion RNA transcript formed by joining exons from two genes (gene A in red and gene B in blue). If either gene A or gene B encodes a lncRNA rather than an mRNA, the resulting fusion RNA is classified as a lncRNA-derived fusion RNA (lnc-fusion). Boxes represent exons, and lines connecting the boxes represent intron regions. The junction within the fusion RNA is termed a breakpoint. (B) Detailed procedure of *LncFusion* for identifying fusions, including both lnc-fusions and mRNA-fusions, from raw RNA-seq data: (1) Integration of Gene Annotations: LncFusion integrates novel lncRNAs identified by the Flnc tool, annotated lncRNAs from the LncBook database, and other annotated genes, including mRNAs from the GENCODE database, to build a comprehensive library of lncRNA and mRNA gene annotations for the targeted biospecimen. (2) Indexing Gene Annotations: LncFusion indexes this lncRNA and mRNA gene annotation library in preparation for breakpoint detection from the RNA-seq data. (3) Breakpoint Detection: LncFusion employs three tools— STAR-Fusion^35^, Arriba^36^, and STAR-SEQR^35^—to detect fusion RNA breakpoints based on the indexed lncRNAs and coding genes (see Fig. 1). These tools were selected for their accuracy and speed in detecting fusions in cancer transcriptomes compared to existing fusion detection tools^47^. (4) Filtering for Confidence: To ensure the reliability of the identified fusions, LncFusion retains only those fusions detected by at least two breakpoint detection tools or fusions with expression levels of FFPM ≥ 0.1 in at least one of the target biospecimens. (5) Further Accuracy Improvements: LncFusion filters out candidate fusions arising from mitochondrial genes, immunoglobulin genes, duplicated genes, paralogues, or fusions involving the same gene, based on the RefSeq curated gene annotation. (6) Final Output: LncFusion outputs the fusion RNAs supported by reads at their breakpoints. This pipeline identifies both lncRNA-derived fusions (including lncRNA-mRNA, mRNA-lncRNA, and lncRNA-lncRNA fusions) and mRNA-derived fusions (i.e., mRNA-mRNA fusions).

Additionally, we have implemented this pipeline into a tool and released it to the GitHub platform (https://github.com/CZhouLab/LncFusion) for the research community. The repository contains source code, sample datasets, and step-by-step instructions for installation and usage.

### *LncFusion* discovers high-confidence lncRNA-derived fusions and mRNA-derived fusions in pediatric neuroblastoma

Neuroblastoma is the most common extracranial solid tumor in children and is often diagnosed at advanced stages, presenting a significant challenge for treatment and survival. To improve the therapy and reduce the burden of long-term side effects in childhood patients, here we first applied *LncFusion* to reveal the lnc-fusions in pediatric neuroblastoma.

Specifically, We applied *LncFusion* to RNA-seq data from 515 neuroblastoma biospecimens across three major cohorts—TARGET, Gabriella Miller Kids First, and St. Jude—and 258 healthy adrenal gland samples from GTEx. This analysis identified 991 high-confidence lnc-fusions and 656 mRNA-fusions (Fig. 2A). By incorporating lncRNAs identified de novo from neuroblastoma and normal adrenal tissues, the pipeline ensured inclusion of cancer-specific and unannotated lncRNAs, expanding beyond conventional gene annotations.

**Figure 2.**
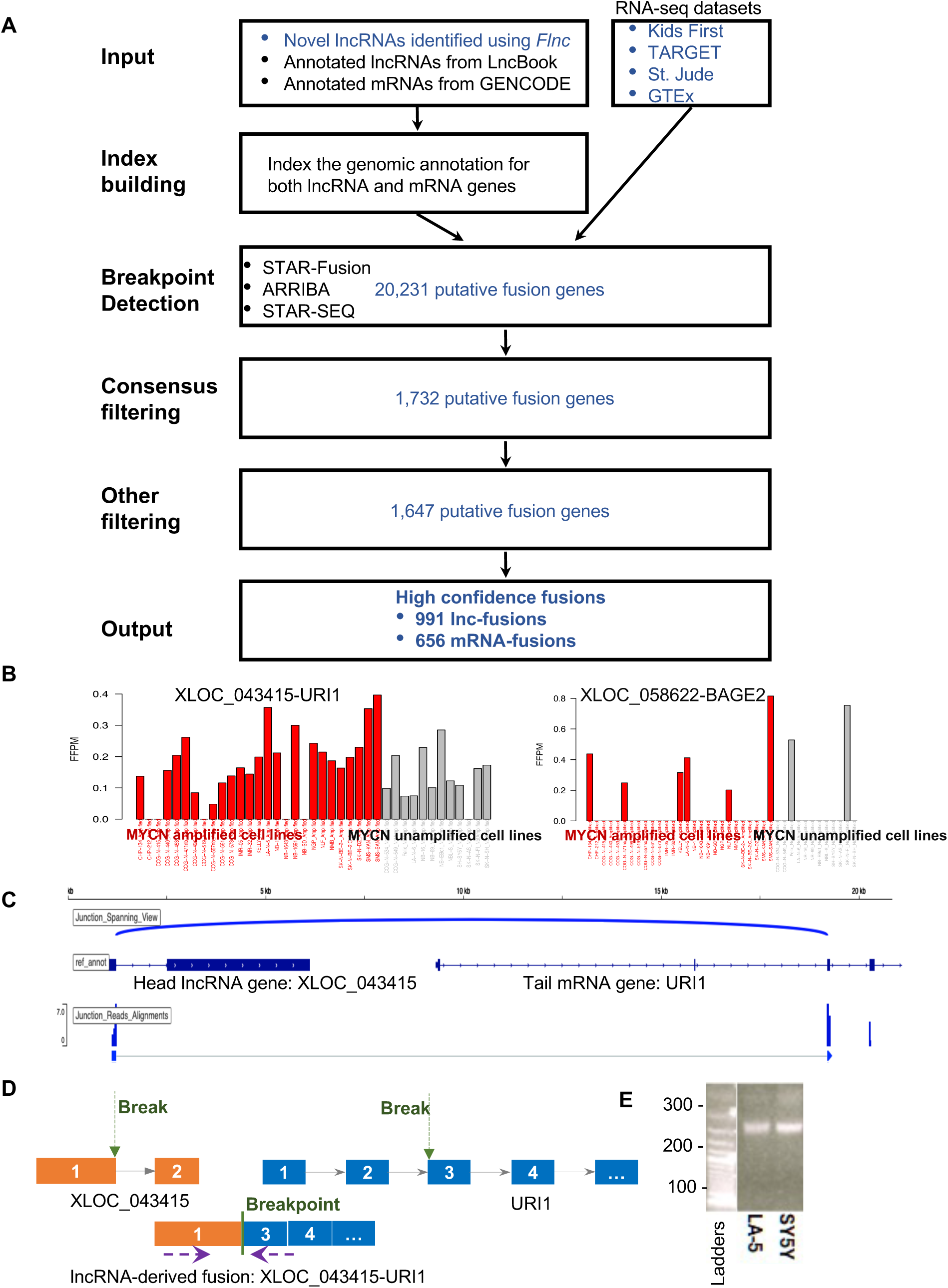
High confidence lncRNA-derived fusions (lnc-fusion) and mRNA-derived fusions (mRNA-fusion) discovered in pediatric neuroblastoma patients. (A) Application of the LncFusion pipeline (described in Fig. 1) to 515 patient samples collected from three cohorts (TARGET, Kids First, and St. Jude), as well as 258 healthy control adrenal gland samples from GTEx. Using Flnc, 4,473 novel lncRNA genes expressed in normal and neuroblastoma adrenal gland biospecimens were identified from the PolyA-selected RNA-seq datasets of the TARGET and GTEx cohorts. In addition to annotated coding and noncoding genes, these adrenal-gland-expressed novel lncRNAs were included in the reference library of gene annotations for fusion detection. Finally, 991 high-confidence lnc-fusions and 656 mRNA-fusions were identified in these pediatric neuroblastoma patients. In this graph, the words in blue represent the detailed steps used by LncFusion to analyze these datasets. (B) Expressions of the identified lnc-fusions were further examined using public RNA-seq datasets of 39 neuroblastoma cell lines to validate the expression of these fusions in independent datasets. As a demonstration, the expression profiles of two lnc-fusions (XLOC_043415-URI1 and XLOC_058622-BAGE2) are shown across these neuroblastoma cell lines. (C) Illustration of the density of spanning reads supporting the breakpoint junctions across the head and tail genes involved in the XLOC_043415-URI1 fusion transcripts. Many spanning reads support the junctions at the 3′ end of the first exon of the XLOC_043415 lncRNA gene and the 5′ end of the third exon of the URI1 coding gene. Additionally, the read densities suggest expression of the first exon of the XLOC_043415 lncRNA gene and the third and downstream exons of the URI1 coding gene, which participate in the fusion transcripts (D-E) RT-PCR validation of identified lnc-fusions in two neuroblastoma cell lines (SH-SY5Y and LA-N-5). (D) Primers (purple arrows) were designed to specifically amplify the spanning junctions across the breakpoints created in the fusions. (E) Two bands amplified by the primers were observed in the gel, consistent in size with the fusion regions in XLOC_043415-URI1. Amplicon size is indicated on the left of the gel.

The initial breakpoint detection step found over 20,000 potential fusions derived from lncRNA or mRNA genes (Fig. 2A), further filtering steps removed the lowly expressed fusions, and these were found by only one breakpoint detection tool to improve the confidence of the identified fusions. This consensus filtering step resulted in about 1,732 potential fusions (Fig. 2A). After removing false positives caused by large gene family and duplicated genes, *LncFusion* found 1,647 high-confidence candidate fusions, including 991 lnc-fusions and 656 mRNA-fusions, in pediatric neuroblastoma.

To validate these findings, we examined the expression of these lnc-fusions in independent neuroblastoma cell line datasets published previously^33^, which includes the RNA-seq data of 39 neuroblastoma cell lines. For example, two representative fusions, XLOC_043415-URI1 and XLOC_058622-BAGE2, were robustly expressed across MYCN-amplified and non-amplified neuroblastoma cell lines (Fig. 2B). These findings demonstrate that the identified lnc-fusions are not artifacts of specific datasets but are consistently expressed in neuroblastoma contexts. The RNA-seq read density of spanning reads, which join the breakpoints of fusions, as well as the density of reads supporting the exons involved in fusion transcripts, were further analyzed and visualized to assess the reliability of the identified fusions. Figure 2C illustrates an example of the raw read density as evidence for the XLOC_043415-URI1 fusion. This fusion is characterized by a junction spanning the 3’ end of the first exon of the lncRNA XLOC_043415 and the 5’ end of the third exon of the coding gene URI1 (Fig 2C and 2D). Notably, the tail gene URI1 is located upstream of the head lncRNA gene XLOC_043415 on chromosome 19. Further, RT-PCR in SH-SY5Y and LA-N-5 cell lines confirmed the breakpoint-specific amplification of the XLOC_043415-URI1 fusion (Fig. 2D-E), providing experimental evidence for its existence.

### lncRNA-derived fusions (lnc-fusions) are widespread in pediatric neuroblastoma

These high-confidence lnc-fusions were found to be widespread and abundant across 515 neuroblastoma biospecimens from three cohorts, outnumbering mRNA-fusions in most individual biospecimens (Fig. 3A). In total, lnc-fusions accounted for approximately 60% of the identified high-confidence fusion events (Fig. 3B). This striking prevalence highlights the under-appreciated significance of lncRNAs in neuroblastoma pathogenesis. Moreover, lnc-fusions exhibited significantly higher expression levels than mRNA-fusions (p < 1.73e-6, Fig. 3C), suggesting that they may have stronger functional roles in pediatric neuroblastoma.

**Figure 3.**
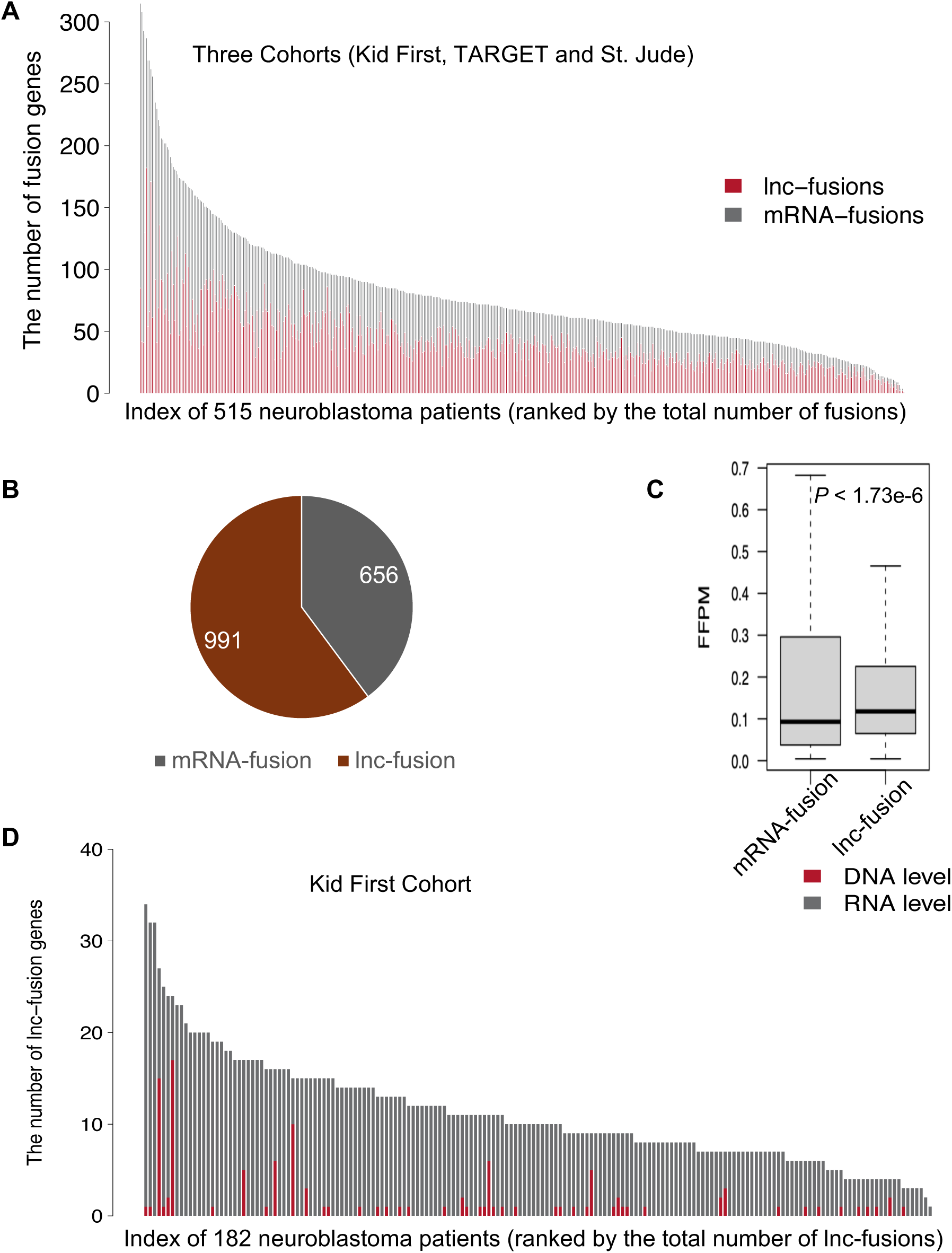
lncRNA-derived fusions (lnc-fusions) are widespread in pediatric neuroblastoma. (A) Distribution of identified high-confidence fusion genes, including lnc-fusions and mRNA-fusions, across 515 neuroblastoma patients from three cohorts: TARGET, Kids First, and St. Jude. In most patient biospecimens, lnc-fusions outnumber mRNA-fusions. All neuroblastoma patients express lnc-fusions. (B) lnc-fusions account for 60% of the identified high-confidence fusions, surpassing the proportion of mRNA-fusions. (C) lnc-fusions generally exhibit higher expression levels than mRNA-fusions, with a statistically significant p-value of <1.73e-6, as determined by the Mann-Whitney Wilcoxon test. (D) Distribution of lnc-fusions formed at the DNA level and RNA level across patients from the Kids First cohort.

Furthermore, we investigated the mechanisms underlying lnc-fusion formation and found that approximately 40% were attributable to chromosomal rearrangements, as evidenced by matched whole-genome sequencing (WGS) data sequenced from the same patient. Whereas the remaining 60% likely resulted from aberrant splicing, which occurs at the transcriptional level without changes to the genomic sequence (Fig. 3D). While most fusion studies have focused on the DNA-level fusions caused by chromosomal rearrangements, our result suggests the potentials of fusions formed purely in the RNA-level. This dual origin underscores the complex genetic and post-transcriptional regulation contributing to neuroblastoma-specific fusion events.

### Tens of lnc-fusions are associated with neuroblastoma and its subtype

To determine whether any lnc-fusions are associated with neuroblastoma or its subtypes, we first clustered all 515 neuroblastoma biospecimens into three major neuroblastoma subtypes: MYCN-amplified, c-MYC-high (highly expressed c-MYC and MYCN-unamplified), and c-MYC-low (low c-MYC expression and MYCN-unamplified) (Fig. 4A-B), following previous studies^48^. This classification reflects differences in prognosis, therapeutic response, and oncogenic pathways among patients, with MYCN-amplified tumors being the most aggressive and c-MYC-low tumors exhibiting least aggressive disease. Most biospecimens were clustered based on their MYCN and c-MYC expression levels using the K-nearest neighbors (KNN) machine-learning method. A subset of biospecimens with annotated MYCN amplification status was used to verify the KNN clustering results, and a few misclassified specimens were manually corrected.

**Figure 4.**
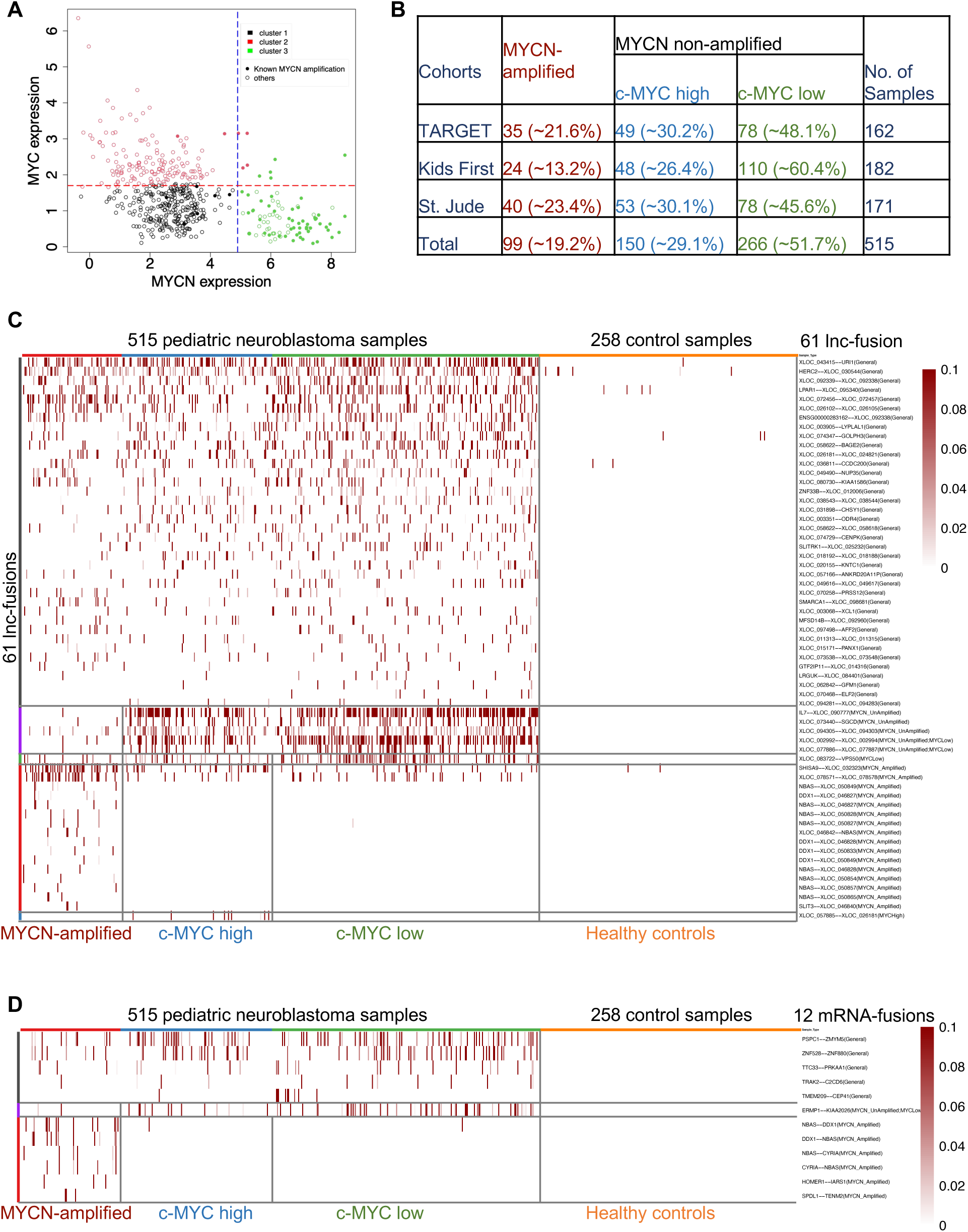
Differentially expression reveals neuroblastoma-associated lnc-fusions. (A) Using the KNN clustering method, all 515 neuroblastoma biospecimens were clustered into three subtypes: MYCN-amplified (cluster 2, in red), C-MYC lowly expressed and MYCN-unamplified (cluster 3, in green), and C-MYC highly expressed and MYCN-unamplified (cluster 1, in black). Each dot represents one biospecimen. The solid dots represent biospecimens with annotated MYCN-amplified or unamplified information, while the hollow dots represent biospecimens with unknown MYCN information that were classified based on the KNN method. (B) The statistical distribution of the three subtypes of neuroblastoma biospecimens across the TARGET, Kids First, and St. Jude cohorts. (C) Expression of 61 neuroblastoma-associated lnc-fusions across neuroblastoma biospecimens and control biospecimens. Among these 61 lnc-fusions, 34 are enriched in all three subtypes of neuroblastoma, while the remaining exhibit subtype-specific expression patterns. (D) Expression of 12 neuroblastoma-associated mRNA-fusions across neuroblastoma biospecimens and control biospecimens. Among these, 5 are enriched in all three subtypes of neuroblastoma, while the others exhibit subtype-specific expression patterns.

Subsequent differential expression analyses identified 61 neuroblastoma-associated lnc-fusions that were either enriched across all neuroblastoma patients or exhibited subtype-specific expression patterns (Fig. 4C), compared to the normal adrenal gland tissues, the primary site of neuroblastoma tumorigenesis. Notably, a greater number of lnc-fusions (61) were associated with neuroblastoma and its subtypes compared to mRNA-fusions associated with neuroblastoma (12 mRNA-fusions) (Fig. 4C-D).

Among these 61 neuroblastoma-associated lnc-fusions, 30 were shared across all subtypes, suggesting general roles in neuroblastoma, while 21 lnc-fusions exhibited subtype-specific expression patterns, being enriched exclusively in MYCN-amplified, c-MYC-high, or c-MYC-low subgroups (Fig. 4C, bottom). For example, XLOC_083722-VPS50 was specific to c-MYC-low patients. These subtype-specific patterns suggest that distinct oncogenic processes drive lnc-fusion expression in these patient subsets, and that these lnc-fusions may have unique oncogenic functions within these subtypes.

### Neuroblastoma associated lnc-fusions might regulate neuroblastoma through several biological processes

Our co-expression analysis revealed that most these Neuroblastoma associated lnc-fusions are significantly co-expressed with multiple known neuroblastoma oncogenes with spearman correlation value greater than 0.3 and p-value <1e-19 across 773 samples (Fig. 5A). These 49 known neuroblastoma oncogenes were listed in the Supplemental Table 1. Specifically, 19 out of 61 neuroblastoma-associated lnc-fusions showed strong co-expression with known neuroblastoma oncogenes, suggesting potential roles in oncogenic pathways (Fig. 5A-B).

**Figure 5.**
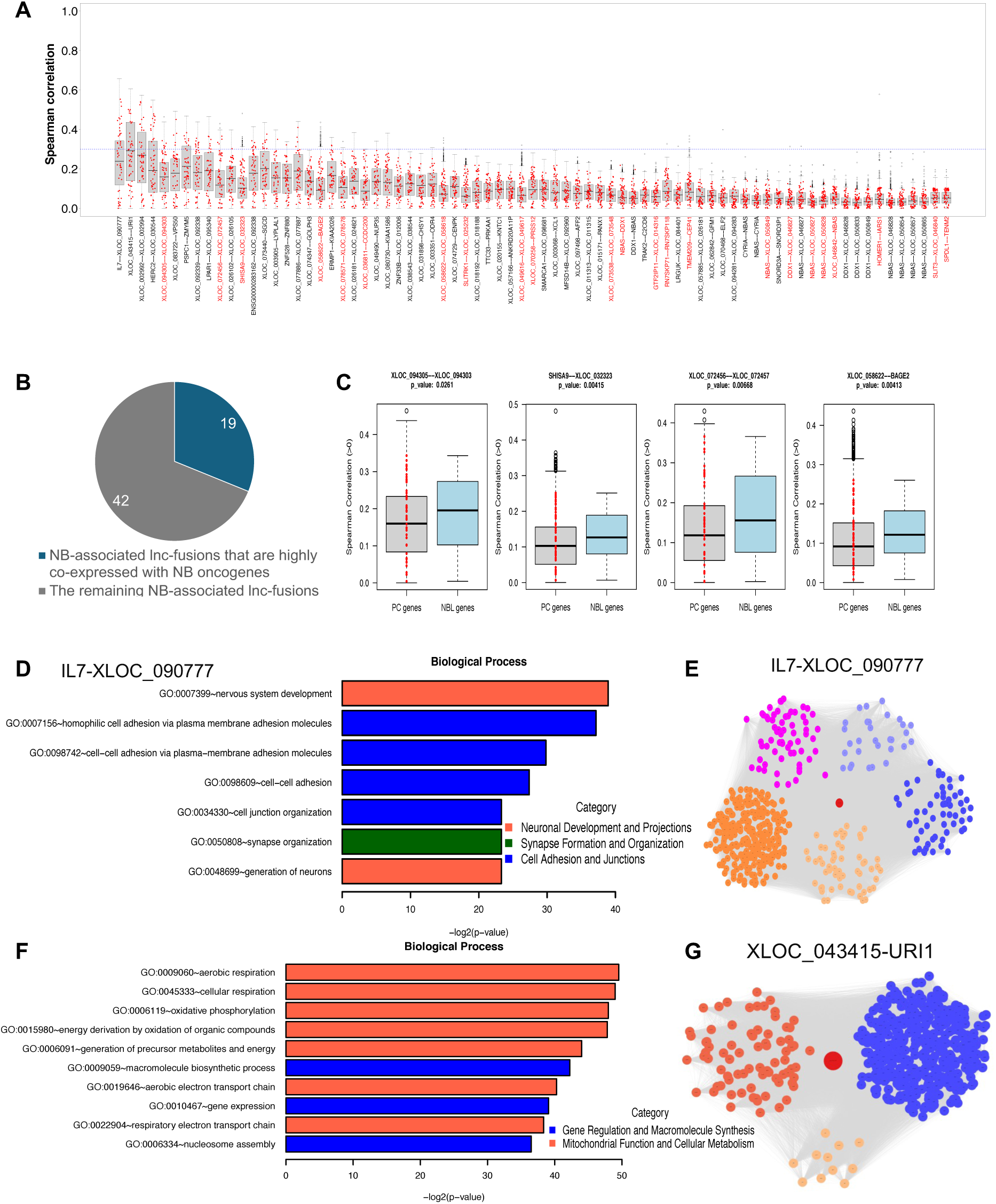
Neuroblastoma-associated lnc-fusions are co-expressed neuroblastoma (NB) oncogenes and form co-expression networks enriched in neuroblastoma-related biological processes. (A) The Spearman correlations of the expression levels of 73 NB-associated fusions with all coding genes, including 61 lnc-fusions and 12 mRNA-fusions. The y-axis represents the Spearman correlation values, and the x-axis lists all 73 fusions ranked by their occurrence frequency in neuroblastoma patients (Fig. 4C and 4D). The red data points represent the correlations of the fusions with any of the 49 known NB oncogenes. The fusions marked in red show co-expression correlations with the 49 NB oncogenes compared to other genes. (B) The pie chart describes how 19 out of 61 NB-associated lnc-fusions exhibit significantly higher co-expression correlations with NB oncogenes compared to other genes. (C) Boxplots displaying examples of lnc-fusions with higher co-expression correlations with NB oncogenes (light blue) compared to other genes (gray). (D) Top Gene Ontology (GO) categories identified for the co-expression clusters containing the IL7-XLOC_090777 fusion. These GO categories are associated with three major biological processes: neuronal development (orange bar), synapse organization (green bar), and cell adhesion (blue bar). The −log10 p-values are shown on the x-axis. (E) Protein-coding genes from co-expression clusters in the top GO categories in Fig. 5D are displayed along with the IL7-XLOC_090777 fusion (red) connected by edges (gray lines). Each circle represents either an mRNA gene or the fusion. Genes involved in neuronal development are labeled in orange, and genes involved in cell adhesion are labeled in blue. Genes in synapse organization belong to either the neuronal development or cell adhesion categories and are labeled in purple to indicate their involvement in multiple biological processes. (F) Top GO categories identified for the co-expression clusters containing the XLOC_090777-URI1 fusion. These GO categories are associated with two major biological processes: gene regulation and macromolecular synthesis (blue bar), and energy metabolism (orange bar). The −log10 p-values are displayed on the x-axis. (G) Protein-coding genes from co-expression clusters in the top GO categories in Fig. 5F are displayed along with the XLOC_090777-URI1 fusion (red) connected by edges (gray lines). Each circle represents either an mRNA gene or the fusion. Genes involved in gene regulation and macromolecular synthesis are labeled in blue, while genes involved in energy metabolism are labeled in orange.

Further Gene ontology (GO) enrichment analysis of co-expression clusters suggested the potential functional roles of lnc-fusions in diverse biological processes. For example, the GO analysis highlighted three major biological processes: neuronal development, synapse organization, and cell adhesion (Fig. 5D)----associated with the IL7-XLOC_090777 fusion. This lnc-fusion is specific to the MYCN-unamplified subtype and is the most frequently occurring lnc-fusion, found in 195 out of 515 neuroblastoma biospecimens (Fig. 4C). It suggests that IL7-XLOC_090777 may participate in neuronal development and synapse-related pathways, implicating its role in neuroblastoma-specific differentiation processes (Fig. 5E).

Similarly, the XLOC_043415-URI1 fusion, the most significant lnc-fusion associated with neuroblastoma across all subtypes (Fig. 4C), was linked to pathways regulating energy metabolism and macromolecular synthesis (Fig. 5F-G). This co-expression analysis suggests its potential roles in neuroblastoma tumor growth and survival through regulating energy metabolism and macromolecular synthesis.

### Several lnc-fusions might be associated with clinical features in pediatric neuroblastoma, suggesting their potential prognostic values

We performed statistical analysis to examine the relationship between neuroblastoma-associated lnc-fusions and multiple clinical features of neuroblastoma, including survival, diagnosis age, severity stage, and risk category. Our survival analyses identified significant prognostic value for specific lnc-fusions. Notably, the HERC2-XLOC_0305044 fusion was associated with poorer survival outcomes in neuroblastoma patients, as demonstrated by Kaplan-Meier analyses (p<0.0001, Fig. 6A). Further subgroup analyses revealed that this fusion correlated with worse outcomes in both MYC-high (p = 0.00074, Fig. 6B) and MYC-low (p < 0.0001, Fig. 6C) subtypes of patients. These findings highlight the potential of HERC2-XLOC_0305044 as a prognostic biomarker.

**Figure 6.**
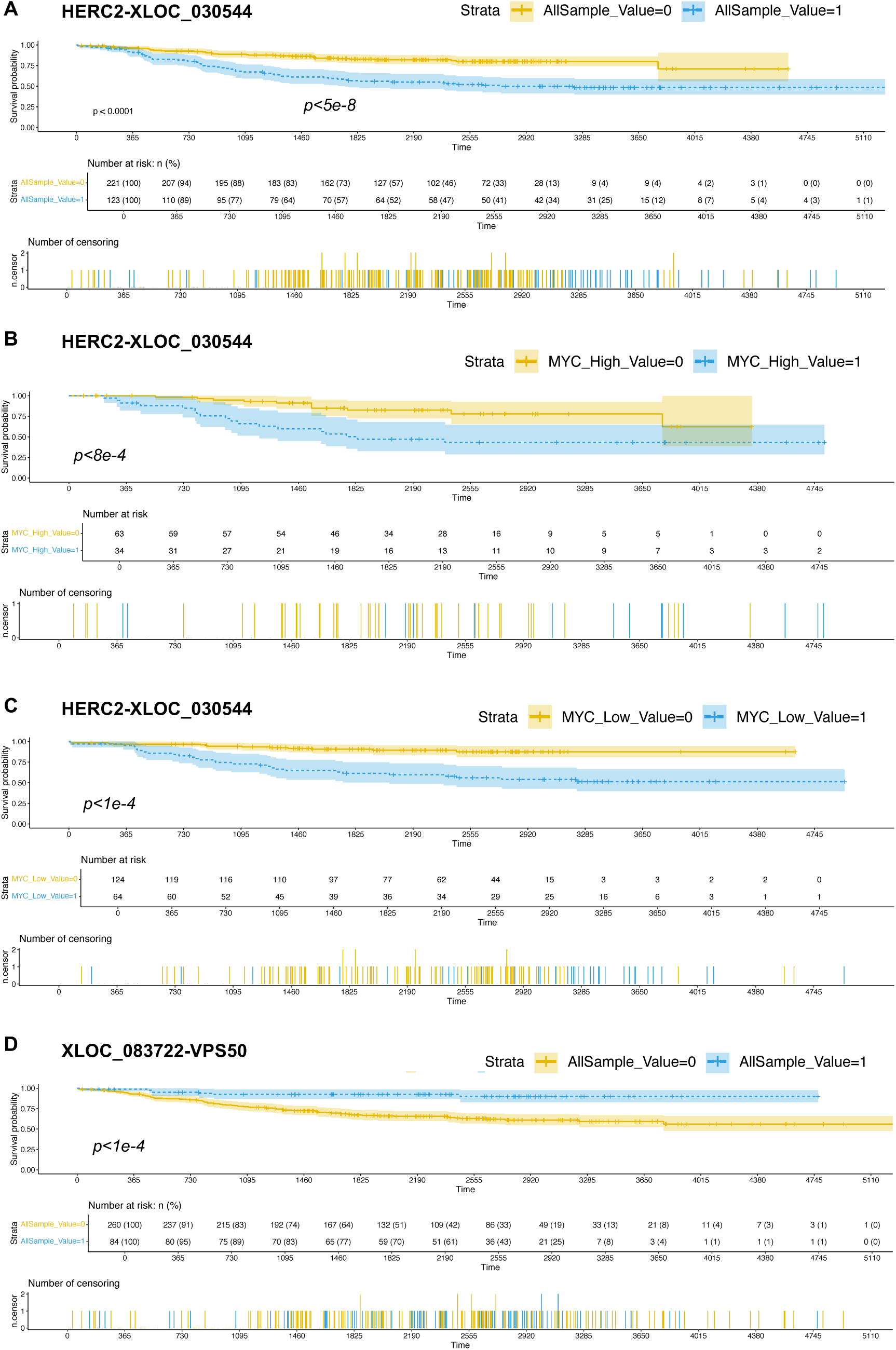
Prognostic significance of the lnc-fusions in neuroblastoma patients. (A) Kaplan-Meier survival analysis for the presence (value=1) or absence (value=0) of the HERC2-XLOC_0305044 fusion among all 221 neuroblastoma patients from the TARGET cohort. (B) Kaplan-Meier survival analysis for the presence (value=1) or absence (value=0) of the HERC2-XLOC_0305044 fusion among 97 neuroblastoma patients with high C-MYC expression and no MYCN amplification (C-MYC high/MYCN-unamplified subtype). (C) Kaplan-Meier survival analysis for the presence (value =1) or absence (value=0) of the HERC2-XLOC_0305044 fusion among 124 neuroblastoma patients with low C-MYC expression and no MYCN amplification (C-MYC low/MYCN-unamplified subtype). Survival differences were assessed using log-rank tests, with p-values shown in each panel. Shaded regions represent the 95% confidence intervals. (D) Kaplan-Meier survival analysis for the presence (value=1) or absence (value=0) of the XLOC_083722-VPS50 fusion among all 221 neuroblastoma patients from the TARGET cohort.

Conversely, several lnc-fusions were associated with favorable outcomes in low-risk patient subsets of neuroblastoma patients. For example, the XLOC_083722-VPS50 fusion was linked to better survival outcomes (Fig. 6D). This association aligns with our observation that XLOC_083722-VPS50 is specifically expressed in the MYC-low subtype, which lacks MYCN amplification and exhibits low c-MYC expression, categorizing it within a generally low-risk group. This suggests that these favorable-associated lnc-fusions predominantly occur in less severe cases, as reflected by the lower proportion of high-risk patients in the cohorts.

This variability in lnc-fusion associations with clinical features indicates that different lnc-fusions may play distinct roles in modulating neuroblastoma aggressiveness. Consequently, these lnc-fusions hold promise as biomarkers for risk stratification, aiding in the prediction of clinical outcomes and the tailoring of therapeutic strategies for pediatric neuroblastoma patients.

## DISCUSSION

In this study, we introduce *LncFusion*, a novel computational pipeline designed to capture both annotated and unannotated lncRNA-derived fusion transcripts (lnc-fusions) alongside mRNA-derived fusions (mRNA-fusions). By integrating multiple fusion-calling tools, stringent filtering steps, and comprehensive lncRNA annotations (including de novo discovery), *LncFusion* addresses critical gaps in existing pipelines, which often overlook non-coding fusion events. This improved workflow ensures the detection of high-confidence lnc-fusions, as evidenced by its application to large-scale pediatric neuroblastoma datasets.

A key finding is the striking abundance and higher expression of lnc-fusions relative to mRNA-fusions in neuroblastoma. These observations underscore the importance of non-coding fusion events in this childhood tumor, highlighting that lncRNAs, previously viewed as purely regulatory, can also form fusion transcripts with potentially far-reaching implications for tumorigenesis. Notably, our data suggest that lnc-fusion formation is mediated by both DNA-level chromosomal rearrangements and RNA-level aberrant splicing, revealing multiple layers of dysregulation. The fact that many lnc-fusions appear to be generated at the transcriptional level suggests they may represent dynamic and potentially reversible oncogenic drivers—an important consideration for future biomarker and therapeutic studies.

Beyond simply expanding the catalog of fusion events, our analysis points to biological relevance. lnc-fusions in neuroblastoma frequently co-occur with established neuroblastoma oncogenes and are associated with pathways linked to tumor growth, metabolism, and neuronal differentiation. These findings imply that lnc-fusions may help orchestrate or fine-tune the oncogenic programs fundamental to neuroblastoma pathology. Moreover, the presence of subtype-specific lnc-fusions, such as those enriched in MYCN-amplified or MYCN-unamplified tumors, suggests a degree of molecular specialization within different patient subgroups. Such subtype-specific fusion events may serve as markers of distinct disease trajectories or therapeutic vulnerabilities.

Our survival analyses underscore the clinical potential of these discoveries. Some lnc-fusions correlate with unfavorable outcomes, pinpointing them as possible indicators of aggressive disease that might be leveraged for early risk stratification. In contrast, others associate with more favorable prognoses, suggesting a biologically protective role, or simply reflecting a less severe disease subtype. Collectively, these data make a compelling case for incorporating lnc-fusion profiling into patient evaluation to refine current stratification models and guide therapeutic strategies.

Despite these advances, there remain challenges and opportunities for future work. The functional mechanisms by which lnc-fusions exert their effects require rigorous in vitro and in vivo validation. Genome editing approaches (e.g., CRISPR/Cas9) and targeted RNA interventions (e.g., antisense oligonucleotides) could illuminate how specific lnc-fusions modulate tumor growth and whether their suppression might convey clinical benefit. Additionally, extending *LncFusion* to other pediatric and adult cancers will determine whether lnc-fusions represent a widespread and underexplored class of oncogenic drivers—one that may inspire new diagnostic and therapeutic avenues.

## CONCLUSION

By unveiling the prevalence, distinct origins, and subtype-specific expression patterns of lnc-fusions in neuroblastoma, our work spotlights lncRNA-derived fusion transcripts as integral components of pediatric tumor biology. The *LncFusion* pipeline, which is publicly available, offers a robust framework for capturing these events and lays the groundwork for future functional and translational studies. Ultimately, the discovery and characterization of lnc-fusions have the potential to refine clinical risk assessment and inform the development of RNA-focused therapies, paving the way toward more personalized treatment for pediatric neuroblastoma and other malignancies.

## SOFTWARE AAVILABILITY

The *LncFusion* software is available on GitHub (https://github.com/CZhouLab/LncFusion) with a tutorial.

## Data Availability

All data produced in the present study are available upon reasonable request to the authors.

## ACKNOWLEDGEMENTS

We thank Dr. Adam C. Resnick and Dr. Jo Lynne Rokita from the University of Pennsylvania for their valuable suggestions regarding the inclusion of the TARGET cohort and neuroblastoma cell line RNA-seq datasets in this project. We also acknowledge the partial assistance of the AI language models ChatGPT-4o and o1-mini in revising the English language of this manuscript.

## FUNDING

This work was supported by NIH R03DE032455-01 (to CZ).

## Authors′ contributions

CZ conceived the study. ZL and CZ, with assistance from JW, designed the study. Computational analyses were conducted by ZL with support from PZ, while JW performed the bench experiments. The results were interpreted by ZL, CZ, and JW. CZ and ZL drafted the manuscript, with input from JW and PZ.

## Conflict of interest statement

The authors have no competing interests to declare.

## SUPPLEMENTAL MATERIALS AND METHODS

### MATERIALS AND METHODS

#### Development of the LncFusion Pipeline

1. **Index Building** We merged three annotation sources—(1) LncBook lncRNAs [citation], (2) GENCODE protein-coding genes (v42) [citation], and (3) novel lncRNAs discovered by Flnc—to create a unified GTF (use.gtf). The reference genome hg38.fa [citation] was used for all alignments.

- **STAR-Fusion Indexing [citation]**

- We built a STAR-Fusion index using perl ctat-genome-lib-builder-master/prep_genome_lib.pl with hg38.fa, use.gtf, and auxiliary databases (Dfam, Pfam, fusion_lib.Mar2021.dat.gz, AnnotFilterRule.pm).
- The final index was generated in STARFusion_Index.
- **ARRIBA Indexing [citation]**

- We generated a STAR index for ARRIBA using hg38.fa and use.gtf with STAR -- runMode genomeGenerate.
- Additional files (blacklist, known fusions, protein domains) were placed in ARRIBA_Index.
- **STAR-SEQR Indexing [citation]**

- We constructed a reference index for STAR-SEQR with the same hg38.fa and use.gtf parameters. The resulting index was stored in STARSEQR_Index.
2. **Breakpoint Detection** Each RNA-seq sample was analyzed with:

- **STAR-Fusion** [citation]
- **ARRIBA** [citation]
- **STAR-SEQR** [citation]

Default or recommended parameters were applied in Singularity containers. Commands included:

- **STAR-Fusion:**

sql
Copy
singularity exec -e $STARFIMG STAR-Fusion \

- -left_fq reads_1.fq.gz \
- -right_fq reads_2.fq.gz \
- -genome_lib_dir STARFusion_Index \
- -output_dir StarFusionOut \
- -CPU 20
- **ARRIBA:**

bash
Copy
singularity exec -B ArribaOut:/output- B ARRIBA_Index:/references:ro \

-B reads_1.fq.gz:/read1.fastq.gz:ro \
-B reads_2.fq.gz:/read2.fastq.gz:ro \
$ARRIBAIMG arriba.sh
- **STAR-SEQR:**

bash
Copy
singularity exec $STARSEQIMG starseqr.py -1 reads_1.fq.gz -2 reads_2.fq.gz \

- m 1 -p StarSeqrOut -t 20 -i STARSEQR_Index \
- g use.gtf -r hg38.fa -vv
3. **Consensus Filtering** We combined outputs from the three callers, retaining fusions detected by ≥2 tools. Fusions unique to STAR-Fusion required FFPM ≥ 0.1. We excluded mitochondrial, immunoglobulin, or highly duplicated genes and removed fusions in which both partners mapped to the same RefSeq transcript.
4. **Output Generation** Fusion events were classified as:

- lnc-fusions: lncRNA–lncRNA, lncRNA–mRNA, or mRNA–lncRNA
- mRNA-fusions: mRNA–mRNA

## Notes

### Competing Interest Statement

The authors have declared no competing interest.

### Author Declarations

dbGaP and GEO databases, and St. Jude Cloud pediatric cancer omics data sharing ecosystem. The St. Jude Cloud pediatric cancer transcriptomics data used in our study consist of individual-level data that had been de-identified prior to their use in this study.

